# Immunomorphogenesis in degenerative disc disease: the role of proinflammatory cytokines and angiogenesis factors

**DOI:** 10.1101/2023.01.05.23284212

**Authors:** N.G. Pravdyuk, A.V. Novikova, N.A. Shostak, A.A. Buianova, O.I. Patsap, A.P. Raksha, V.T. Timofeyev, V.M. Feniksov, D.A. Nikolayev, I.V. Senko

## Abstract

Back pain (BP) due to degenerative spinal injury is a severe, often disabling disease and it tends to decrease age of onset. The cellular and molecular basis of the disc and adjacent structures degradation remains in the focus of pathophysiologists and pathomorphologists close attention. The aim of the study was to determine the expression level of proinflammatory cytokines (IL-1β, IL-6, IL-17) and angiogenesis markers (VEGF-A and CD31) in intervertebral disc (IVD) tissue, their association between each other and between disc degeneration in young people with discogenic BP. In patients who underwent discectomy for a disc herniation, a clinical examination, magnetic resonance imaging (MRI) of the lumbar spine, histological and immunohistochemical analyses of these factors in the disc tissue were performed in comparison with the parameters of healthy group samples (controls). Histology image analysis of IVD fragments of the degenerative disc disease (DDD) group detected zones of inflammatory infiltration, combined with vascularization, the presence of granulation tissue and clusters of chondrocytes in the tissue of nucleus pulposus (NP). Statistically significant expression of IL-1β, IL-6, IL-17 and VEGF-A (especially on the chondrocytes surface) and CD31 (in the disc matrix) was evident in the samples of the DDD group compared with the controls (p < 0.0001), that showed a strong correlation with the histological disc degeneration stage (r > 0.5; p < 0.0001). This denotes a high immunoinflammatory potential of chondrocytes and demonstrates their altered morphogenetic properties. The coincidence of the spatial expression of IL-1β and IL-17 in the perivascular zone endothelium and in the vascular lumen (p < 0.01) was found, which point at the inflammatory synergy of these cytokines. High expression of VEGF-A prevailed on the surface of chondrocytes in cell clusters compared to the matrix (p < 0.0001), indicating that the NP cells trigger the angiogenesis. The absence of CD31 in the cracks of NP with high expression in the disc matrix states the secondary nature of IVD defects due to degeneration, and not due to vascular ingrowth. A high number of patients with Modic changes according to MRI data shown the contribution of cytokines to the formation of reactive spondylitis and the clinical course of BP. These results obtained will help to development molecular/cellular targets and basic strategies for therapy of BP with DDD in the early stages in young people.

## INTRODUCTION

The overwhelming number of back pain (BP) cases is due to a degenerative lesion of the spine, which is accompanied by a complex of changes in the vertebral-motor segment. Modern research in the pathogenesis of spinal degeneration covers all the proposed mechanisms: from molecular/cellular and genetic to psychosocial, which leads to the formation of a multimodal treatment approach for BP, especially during the BP chronization.

Over the past two decades the study of degeneration of the intervertebral disc (IVD) and adjacent vertebral bodies at the microlevel has focused the attention of researchers on the immunological mechanisms of cellular matrix damage. Thus, it was shown that in the early stages of disc degeneration, there is a shift in the balance of anabolic and catabolic activity of extracellular matrix molecules and IVD chondrocytes, which occurs with the direct participation of proinflammatory cytokines. The early stage of disc degeneration – preclinical – consists in the disintegration of the intercellular substance, hypoxemia, acidification of the biochemical medium and accumulation of lactate [1], loss of hygroscopicity of glycosaminoglycans and chondroitin sulfates of the nucleus pulposus (NP) [2]. Fragmentation products include hyaluronic acid, biglican, versican, decorin, elastin and laminin, which aggravate the inflammatory cascade in the disc [3]. Moreover, inflammatory cytokines induce the expression of matrix-destroying proteins [4]. This leads to a decrease in the elasticity and surface tension of the disk NP with an increase in the recovery time of its height under axial loads, dehydration, and violation of the cellularity of already extremely small chondrocytes [5]. Induction of inflammatory activity in the disc and in adjacent structures leads to advanced stages of degeneration, protrusions, hernias and stenosis of the spinal canal [6].

Traditional inflammation, as is known, involves the formation of granulation tissue consisting of macrophages and T-lymphocytes. Previous studies have shown that pro-inflammatory molecules involved in the development of spinal degeneration include IL-1, IL-2, IL-4, IL-10, IL-12, IL-15, IL-17, PGE2, IFN-γ, whose expression increases locally, as well as IL-6, IL-8, TNF-α, the expression of which increases in the blood serum [3]. It is known that macrophages are the most numerous of all inflammatory cells. After activation, they secrete cytokines and compounds that cause disc degeneration, and can even spread to nearby intact disc tissues in advanced stages of degenerative disc disease (DDD) [7]. This implies an active migration of immunocytes to the lesion/inflammation site along the vascular bed. However, the initial signs of degeneration are detected in discs that have retained their anatomical integrity, i.e. in the absence of obvious mechanical micro- and macro-injuries that could act as a “window of migration” of immunocompetent cells with the launch of inflammation. In view of this, it remains unclear whether macrophages play a dominant role in disc degeneration by analogy with other tissues, or the initiation of inflammation begins in their absence due to the transformation of the cells of the NP and the annulus fibrosus (AF) themselves, and what is the role of disc vascularization with loss of its immunological tolerance and repair. It is likely that inflammatory cytokines control the degenerative process in the disc, participate in the mechanisms of pain and are the most relevant target for pathogenetic therapy [8].

The aim of our study was to identify the expression level of proinflammatory cytokines (IL-1β, IL-6, IL-17) and markers of angiogenesis (VEGF-A and CD31) in IVD tissue and their correlation between tissue degeneration and indicators of discogenic BP in young people.

## MATERIALS AND METHODS

### Patients

The study was approved by the Ethics Committee of Pirogov Russian National Research Medical University (protocol № 13 dated November 30, 2020). 34 patients (17 men and 17 women) were sequentially selected in the neurosurgical department of the N.I. State Clinical Hospital No. 1. Pirogov. They were scheduled to have a microdiscectomy due to progressive BP and/or neurological symptoms associated with a hernia. The inclusion criteria in the DDD group were: young age (median age – 32.00 years [29.00–39.50]) and BP associated with IVD hernia confirmed by instrumental data (magnetic resonance imaging (MRI)). Exclusion criteria were: spinal injury at the time of the study and in the anamnesis, tumor, infections of the spine and other organs, inflammatory spondyloarthritis, surgical interventions in the previous 30 days. The intensity of BP was clinically assessed according to VAS: the types of pain were acute or chronic (according to the classification of The International Association for the Study of Pain (IAPS) 2021), the pain lasts years (both persistent and recurrent) and the functional limitation of spinal mobility was scored by the Backache-Index. All patients underwent MRI on a Toshiba1.5 tesla device in three sections (sagittal, coronary and axial).

### Tissue samples

There were obtained 34 samples of disc tissue from patients of the DDD group during the discectomy and 7 samples of control disc tissue. The biopsy specimen consisted of multiple fragments of whitish, dense elastic tissue and cartilaginous density tissue with a total size of 6×5×1 cm. Control samples were obtained during spinal stabilization and removal of the dropped segments of intervertebral cartilage L4/L5, L5/S1 in 7 healthy individuals due to acute auto injury within 1 hour after the incident. The age of the control group varied from 27 to 44 years. The patients had no history of diseases of the spine and musculoskeletal system, infectious or oncological diseases, and did not abuse narcotic drugs. Patients of both groups signed an informed consent to participate in the study.

### Histological examination

IVD tissue samples were fixed immediately after discectomy in 4% paraformaldehyde/phosphate-buffered saline (pH = 7.4) for 72 hours in room temperature. In the presence of calcification or residual bone material, decalcification in HCl was performed, then samples were processed into paraffin wax. Four-micron sections from the tissue blocks were cut and stained with hematoxylin and eosin according to standard protocol.

### Immunohistochemical (IHC) analysis

All sections were examined by IHC on the automated VENTANA BenchMark ULTRA platform. For investigating angiogenesis, a primary mouse anti-VEGF-A (1:500 dilution; Cloud-Clone Corp., USA) and primary rabbit anti-CD31 (1:150 dilution; ab28364, Abcam, Cambridge, UK) were used. To identify processes of inflammation, were also applied primary mouse anti-IL-1β, anti-IL-6 and anti-IL-17 (1:150 dilution; Cloud-Clone Corp., USA).

### Morphometric analysis

All slides were visualised using a Axio Imager.Z2 microscope (Carl Zeiss, Germany). Cell counts were performed on all independent fields of photomicrographs captured with EC Plan-Neofluar 40X objective. The region imaged was nucleus pulposus. Positive signal was observed as dark brown/yellow brown coloring. Image analysis was carried out in the Fiji program. The stage of degeneration was established in accordance with the criteria of Sive [9], where 0-3 points were considered as the norm, 4-9 – degeneration, and 10-12 – severe degeneration. The markers expression was assessed by percentage ratio of immunopositive cells to the total number of cells.

### MRI data analysis of the lumbar spine

According to the MRI data, the IVD height was assessed at the hernia level, the Pfirrmann DDD stage at the operated level (disappearance of the difference between NP and AF and a decrease in the intensity of the signal from the NP), the presence of reactive changes in the adjacent vertebral bodies according to the Modic type (1, 2, 3 types) – changes in the MR signal intensity from the bone marrow in the T2-weighted image and with fat suppression (STIR), the severity of the lesion Modic-changes in area in the sagittal and/or coronary section and the presence of an erosive type of lesion of the vertebral body endplates (EP).

### Statistical analysis

Statistical data was analyzed using Graph Prism 8.0.1 (GraphPad, La Jolla, CA, USA). The distributions were determined to be parametric by Shapiro-Wilk testing. Data with a normal distribution were presented as the values of the mean ± SD, non-normal variables were presented as median [Q1, Q3]. To verify the differences between the expression of immunohistochemical markers and the severity of histological degeneration, Mann-Whitney U test was used. Spearman’s criterion was used for correlation analysis. A p-value < 0.05 was considered statistically significant.

## RESULTS

### Clinical characteristics of patients and MRI data

BP characteristics in the DDD group are presented in table 1. Pain intensity median corresponded to 62 [46.75–87.00] mm. Most of the patients (30 or 88.24%) suffered from chronic BP. At the same time, the recurrent type of pain was found in 73.53%, and persistent (permanent without a *bright* gaps) – in 26.47%. The duration of the pain syndrome ranged from 0.1 to 25 years. 28 of patients (82.35%) had moderate and severe degrees of functional disorders according to the BAI index.

**Table 1.**
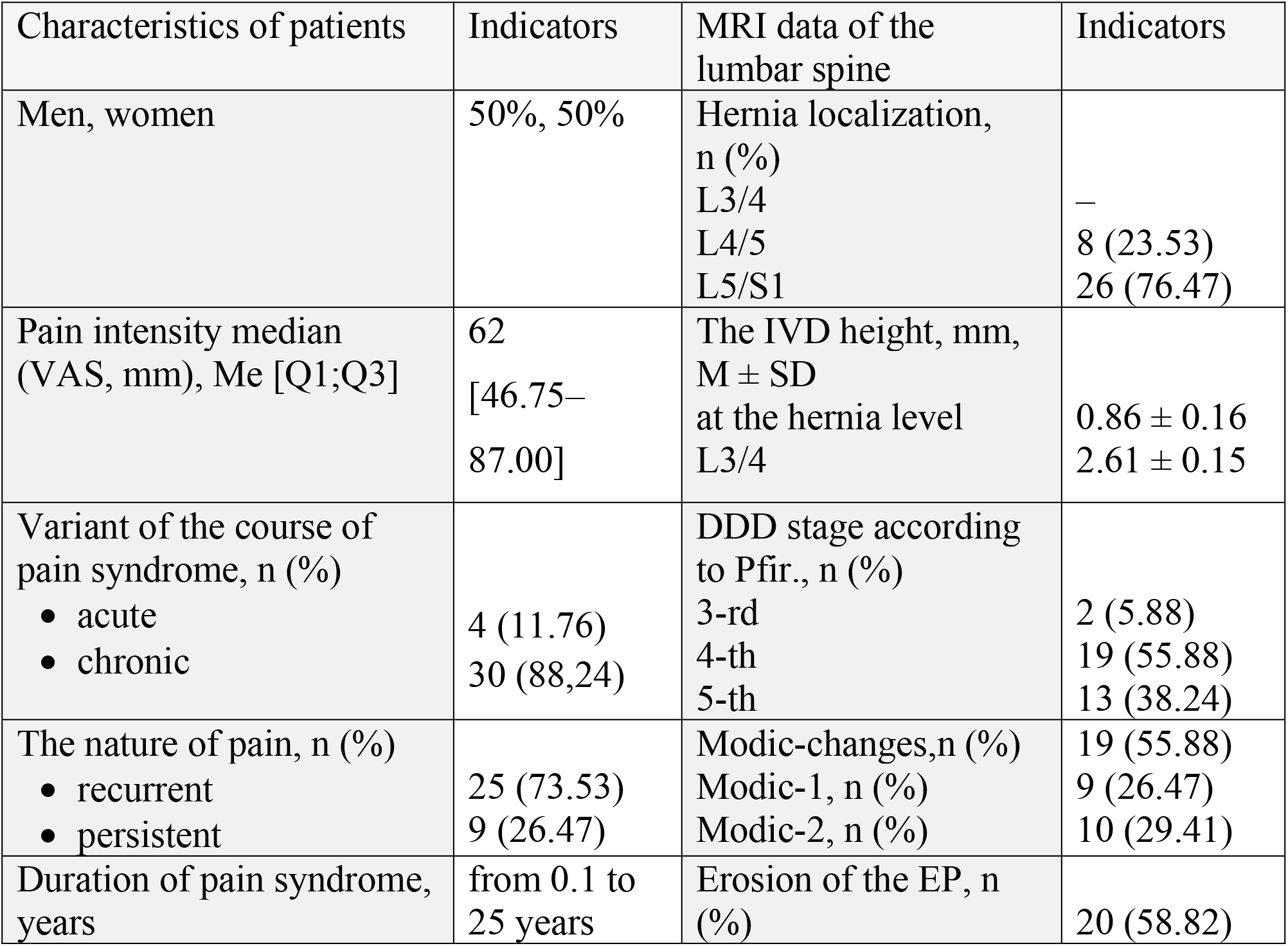
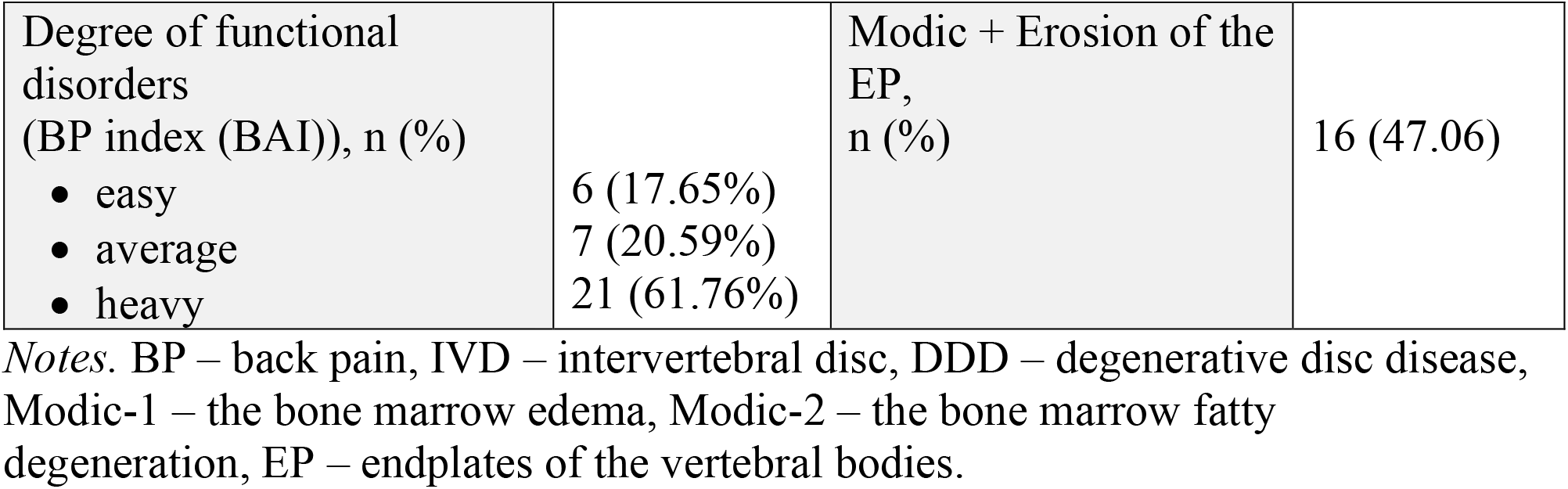
Clinical and instrumental indicators in the DDD group, n=34.

**Table 2.** Indicators of the expression level of the studied markers in patients determined by the IHC.

| Marker | Expression level in patients, average value %, n=34 | Expression level in controls, n=7, average value % |
| --- | --- | --- |
| **IL-1β** | Diffuse in the matrix: $13.06 \pm 3.65$<br>(around the vessels $79.24 \pm 8.12$)<br>In cell clusters: $56.15 \pm 24.40$ | Diffuse in the matrix: $1.17 \pm 0.47$ (around the vessels $4.18 \pm 1.72$)<br>In chondroblasts, the NP is $16.43 \pm 10.10$ |
| **IL-6** | Diffuse in the matrix: $16.33 \pm 5.40$<br>In cell clusters: $49.65 \pm 21.41$ | Diffuse in the matrix: $3.93 \pm 1.17$<br>In chondroblasts, the NP is $29.86 \pm 8.45$ |
| **IL-17** | Diffuse in the matrix: $14.15 \pm 5.12$<br>(around the vessels $51.89 \pm 7.35$)<br>In cell clusters: $63.71 \pm 21.12$ | Diffuse in the matrix: $0.57 \pm 0.35$ (around the vessels $3.51 \pm 1.60$)<br>In chondroblasts, the NP is $8.71 \pm 5.77$ |
| **VEGF-A** | Diffuse in the matrix: $16.82 \pm 5.70$<br>In cell clusters: $74.68 \pm 19.17$ | Diffuse in the matrix: $0.51 \pm 0.12$<br>In chondroblasts, the NP is $17.43 \pm 7.74$ |
| **CD31** | Diffuse in the matrix: $19.71 \pm 9.25$ | Diffuse in the matrix: 0 |

### Results of MRI of the lumbar spine

Localization of IVD hernia in 23.53% (n=8) of individuals was at the level of L4/5, in 76.47% (n=26) – at L5/S1. The average value of the IVD height demonstrated a significant narrowing of the intervertebral space at the level of the operated disc (L4/5, L5/S1) compared to the L3/4 level – the average values of the IVD height were 0.86 ± 0.16 and 2.61 ± 0.15 mm, respectively (p < 0.01). The stage of IVD degeneration at the discectomy level according to Pfirr. corresponded to the 4th and 5th stages in 55.88% (n=19) and 38.24% (n=13), respectively. In half of the patients (n=19; 55.88%), reactive changes in the adjacent vertebrae bodies were detected: the bone marrow edema (Modic-1) in 26.47% (9 patients) and fatty degeneration of the bone marrow (Modic-2) in 29.41% (10 patients) (fig 1). Almost half of the patients in the DDD group (n=16; 47.06%) had a combination of IVD hernia plus Modic-changes with EP erosive lesion of the adjacent vertebral bodies.

**Figure 1.**
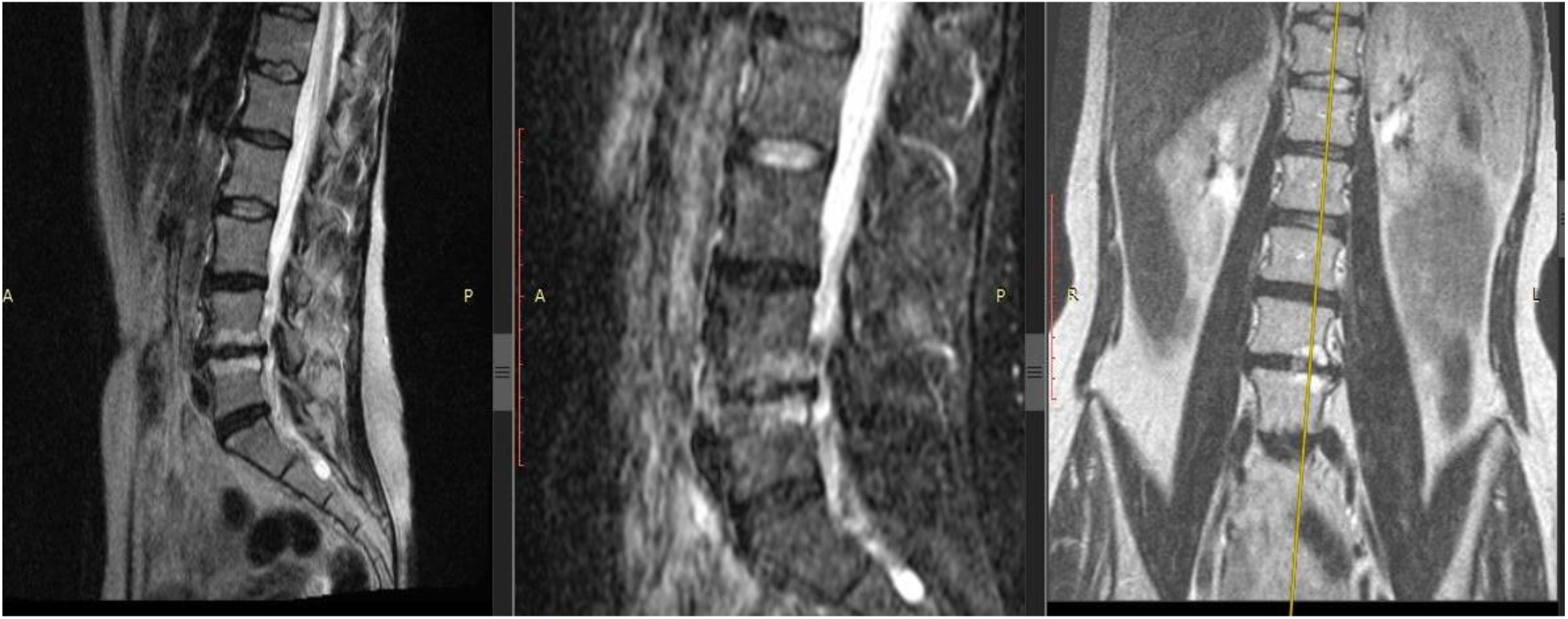
Patient with BP. MRI of the lumbar spine, modes T2- and T2-weighted image with fat suppression in sagittal sections (left and center images, respectively), T2-weighted image in the coronary section (right image). Right-sided lumbar scoliosis, the 5th stage of DDD by Pfirrmann at the L4/5 and 4th stage – at the L5/S1 level, with IVD hernias L4/5 and L5/S1, erosion of the endplates and Modic-changes type 1 in the vertebral bodies L4/5 (yellow axis carried out via Modic-1).

### Histological and morphometric data

In the IVD samples of the DDD group, inflammatory infiltration zones were identified, combined with vascularization (fig 2A) and the presence of granulation tissue. There were also the regions of hyaline cartilage with inflammatory cells infiltration.

**Figure 2.**
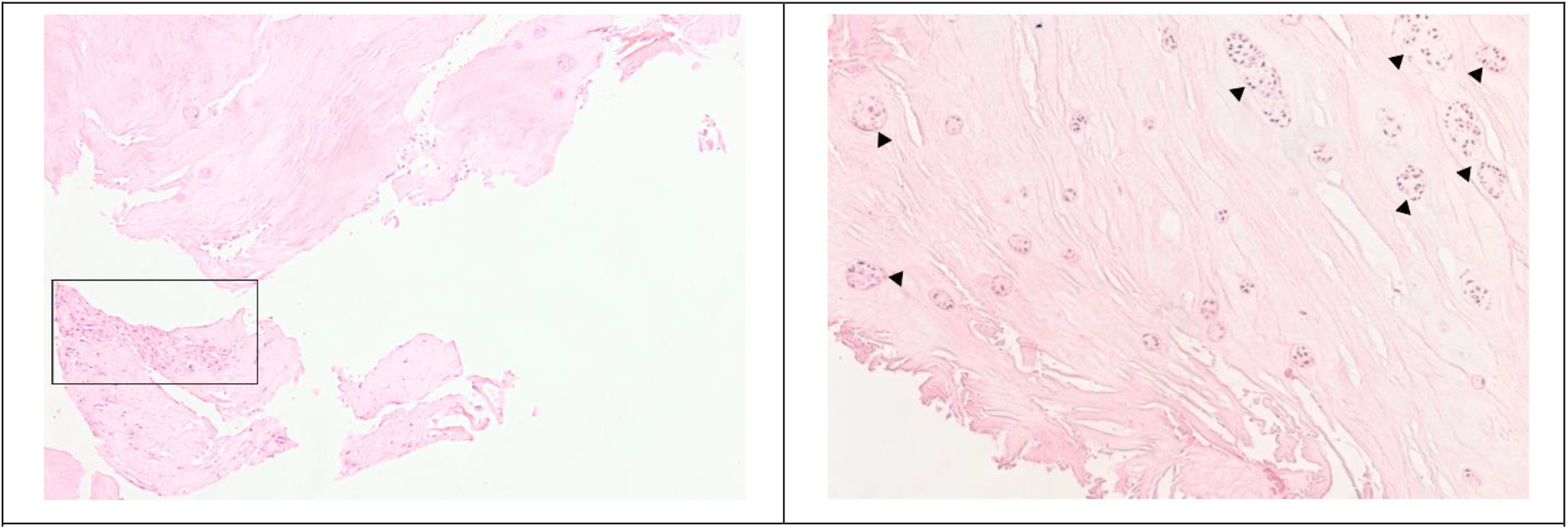
IVD sample obtained as a result of microdiscectomy in the patient in her 40s, with a hernia of the L5/S1 vertebrae. Own data, 2020. Hematoxylin-eosin staining. A. Light microscopy, magnification x100. Zones of inflammatory infiltration, combined with vascularization and granulation tissue. B. The sample of the same patient, magnification x100. Clusters of NP cells (triangles) are a characteristic sign of disc degeneration.

The morphological degeneration stage in the samples of the DDD group was Me = 1,000 [0.000–3.000] points, while of the control group was Me = 7.000 [6.000– 10.000] (p < 0.0001). In the samples of DDD patients, clusters of NP cells (clusters of chondrocytes) were detected (fig 2B), especially in damaged areas that were absent in the control samples, which can be regarded as another morphological sign of disc degeneration. The average area of chondrocyte cluster was 3645.393 ± 551.701 microns^2^ and the average number of cells in them was 9.411 ± 3.382.

### Immunohistochemical study

Immunohistochemical staining revealed a statistically significant difference in the expression levels of all the proinflammatory cytokines and vascular endothelial growth factors determined in all patients with BP and DDD compared with the control group (fig 3 and 4, tab 2).

**Figure 3.**
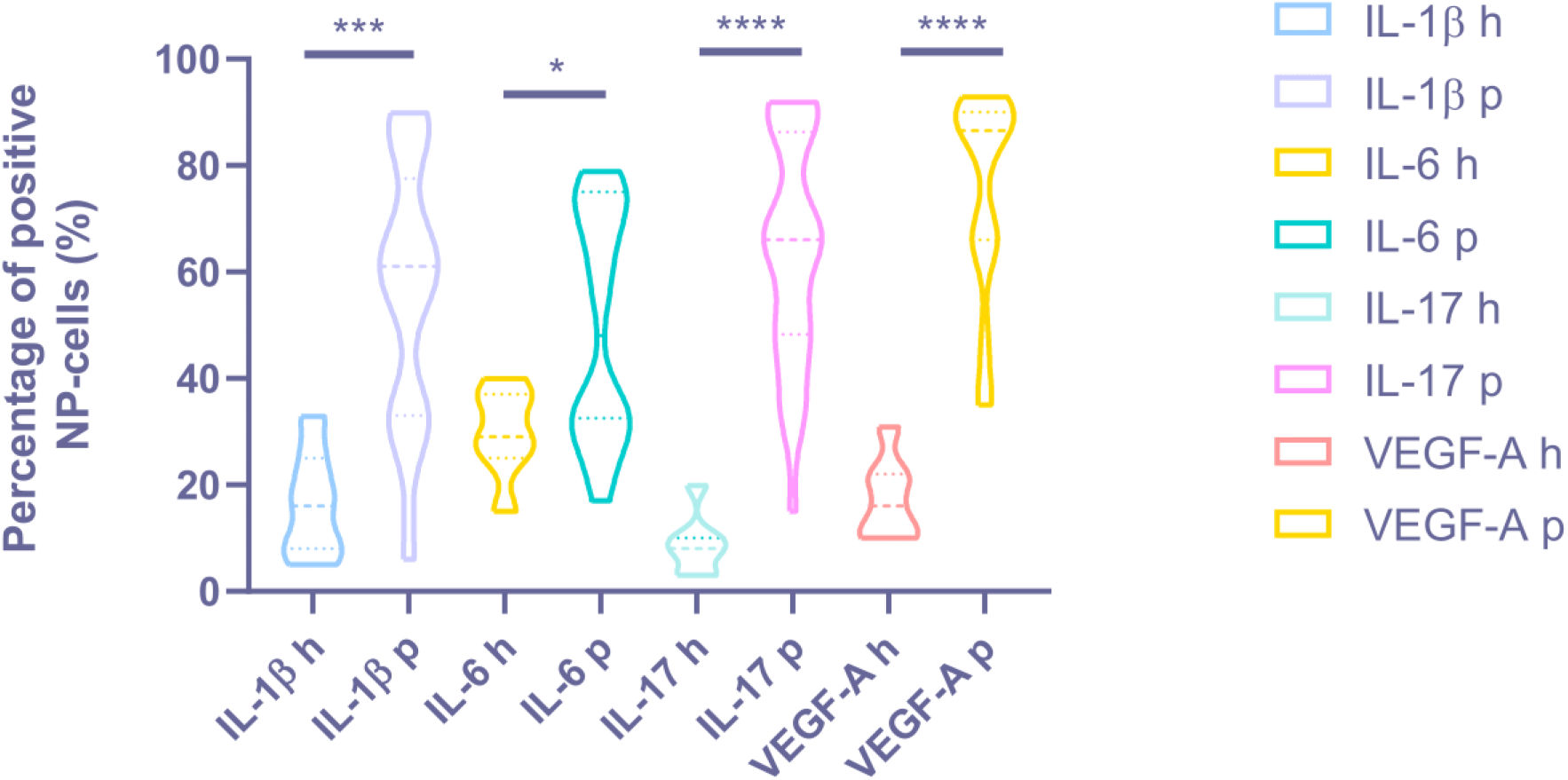
The percentage of positive NP-cells in the IVD tissue of the DDD (“p” – patients) and control group (“h” – healthy) discs. * p < 0.05. *** p < 0.001. **** p < 0.0001.

**Figure 4.**
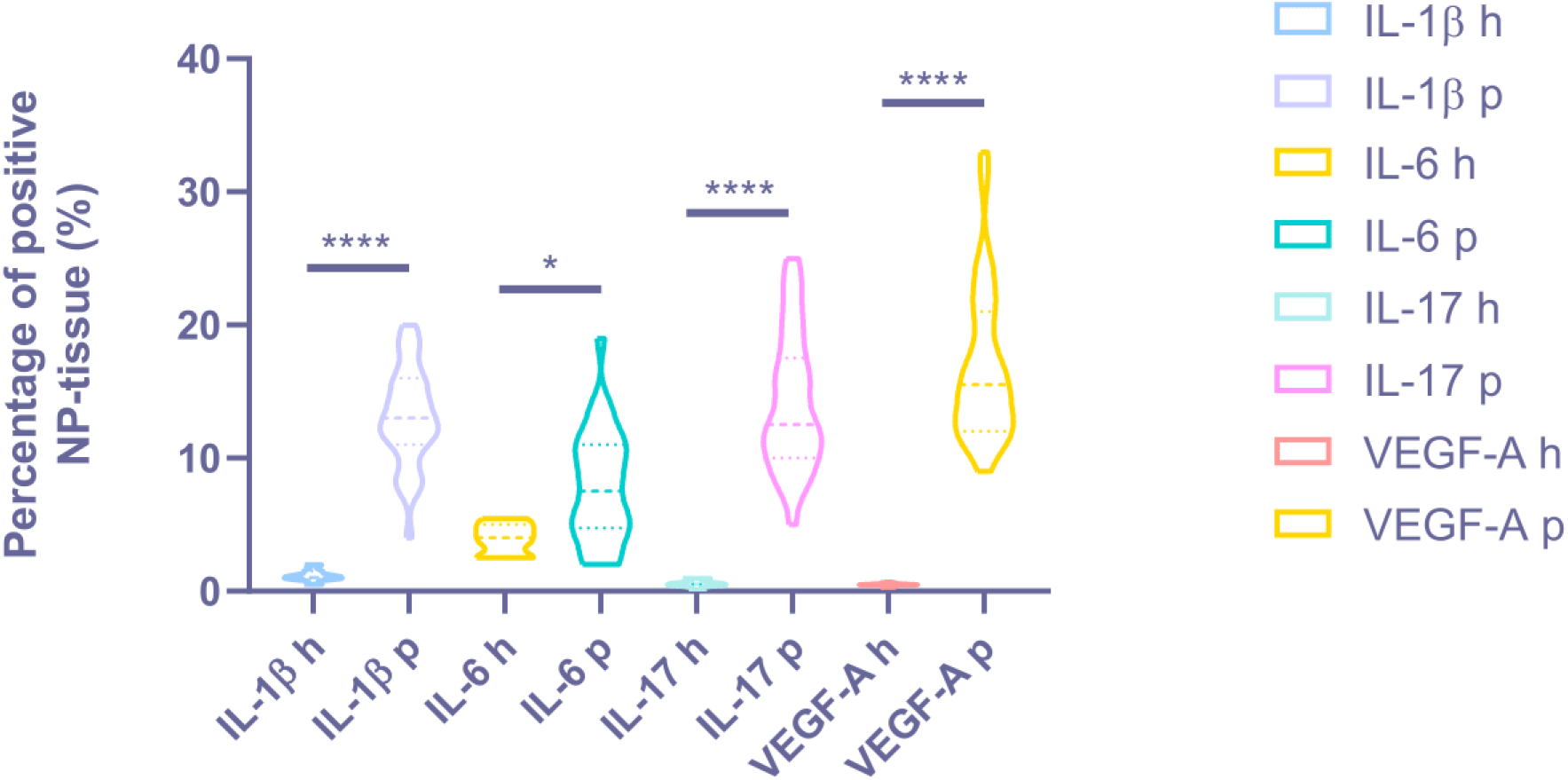
The cytokine expression degree in the IVD matrix of the DDD (“p” – patients) and control groups (“h” – healthy). * p < 0.05. *** p < 0.001. **** p < 0.0001.

**Figure 4.**
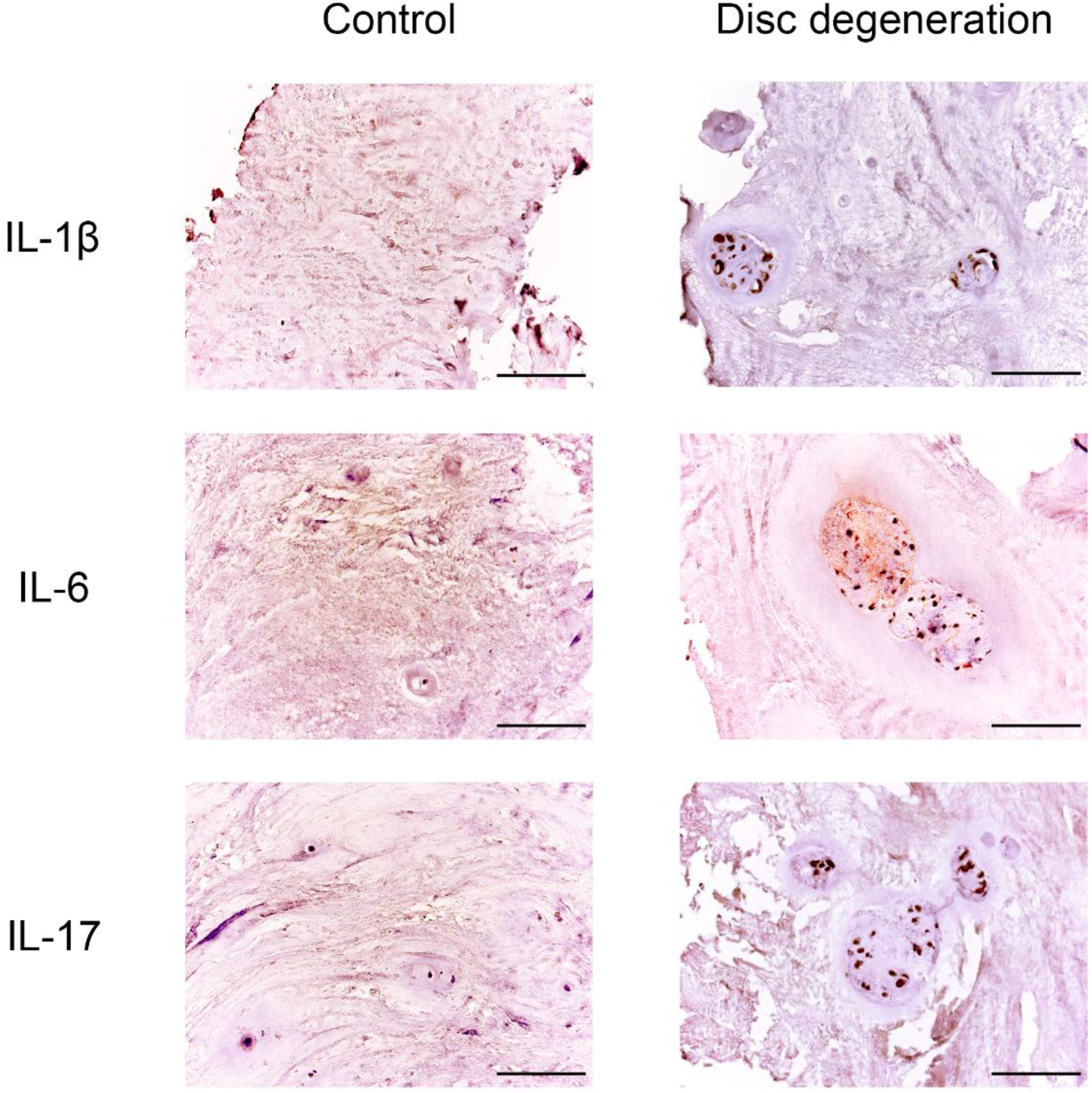
Expression of IL-1β, IL-6, IL-17 in IVD tissue. Scale bars: 100 μm.

High expression IL-1β level was revealed in the NP chondrocytes (DDD: 56.15 ± 24.40%, control: 16.43 ± 10.10%; p < 0.001) and around the vessels; medium expression – in the disc matrix (p < 0.05) (Figure 4). Expression of IL-1β was positively correlated with histological degeneration stage (r = 0.606, p < 0.0001). IL-1β was expressed in two samples of the control group.

The IL-6 expression was medium in the chondrocytes of the NP (DDD: 49.65 ± 21.41%, control: 29.86 ± 8.45%; p < 0.05) (fig 4). The correlation coefficient of 0.597 (p < 0.0001) indicated a moderate positive correlation between IL-6 expression and histological degeneration stage.

High IL-17 expression in NP cells (DDD: 63.71 ± 21.12%, control: 8.71 ± 5.77%; p < 0.0001) and around vessels (fig 4) states the appearance of this cytokine due to activation of NP chondrocytes and probably due to the migration of immune cells through newly formed disk vessels. In the DDD group, a coincidence of the spatial expression of IL-1β and IL-17 was found (in 21.62 ± 9.90% of immunopositive chondroblasts) – in the endothelium and in the perivascular zone, as well as in the vascular lumen (p < 0.01). It confirms the hypothesis that they act synergistically during inflammation processes. The positive correlation between IL-17 expression and histological degeneration stage was strong (r = 0.616, p < 0.0001). This marker was expressed around the vessels in 2 control samples, along the disc edges.

High expression of VEGF-A was detected in clusters of cells (it occupies more than 3/4 of the cluster) along with weak expression in the matrix and around single cells of NP (DDD: 74.68 ± 19.17%, control: 17.43 ± 7.74%; p < 0.0001) (fig 5), indicating that NP cells (as single, and in clusters) trigger the angiogenesis process. The moderate correlation between VEGF-A expression and histological degeneration stage was positive (r = 0.519, p = 0.001).

**Figure 5.**
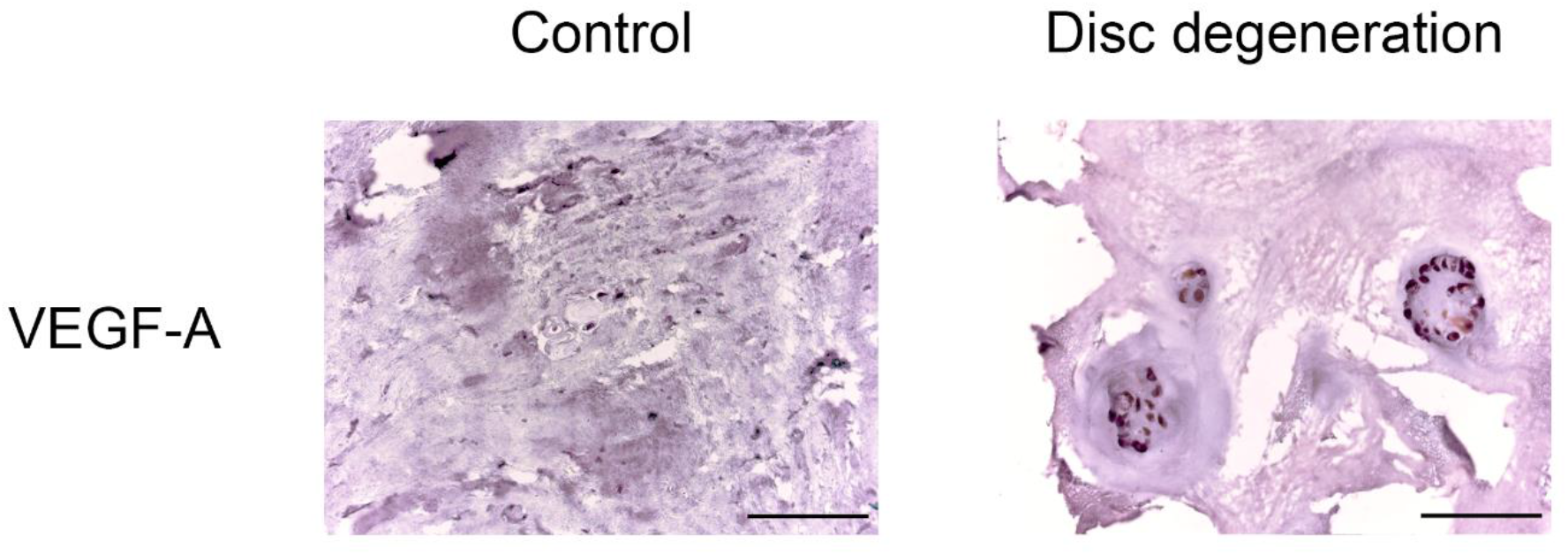
VEGF-A expression in IVD tissue. Scale bars: 100 μm.

CD31 was found in 22 patients. High expression of CD31 was detected around NP single cells and in the disc matrix, where were no vessel (fig 6). This marker was also presented in the endothelium of the vessels in granulation tissue in IVD samples of 4 patients. Cracks in the disc were not stained on CD31, which excludes the primary role of vascular ingrowth in the formation of IVD defects. CD31 was not expressed in any control sample.

**Figure 6.**
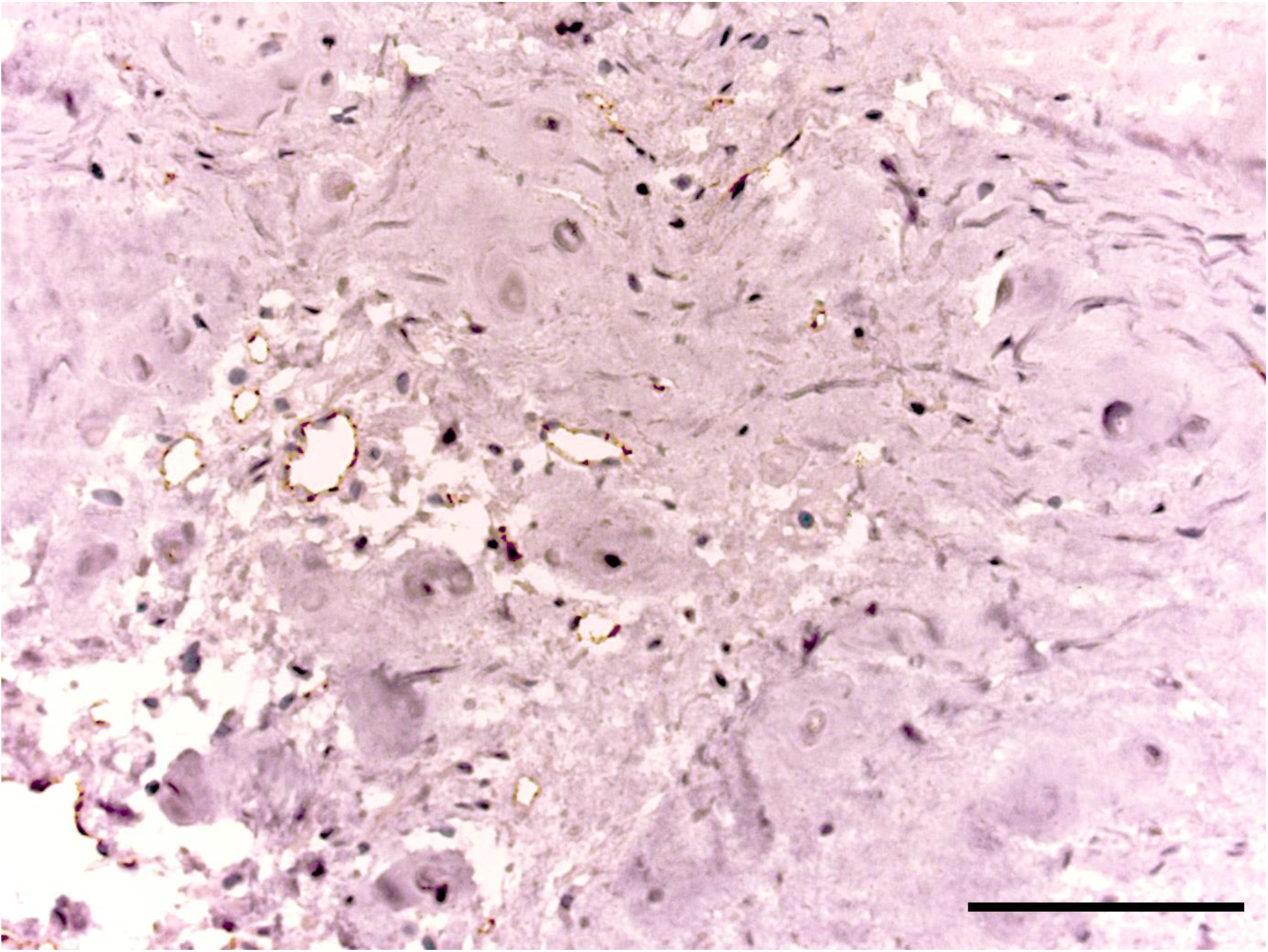
CD31 expression in the IVD tissue of the patient of the DDD group. Scale bar: 100 μm.

## DISCUSSION

All patients were young and, regardless of gender, had intense pain syndrome associated with severe IVD degeneration according to MRI (4th and 5th stages of DDD according to Pfirrmann), a long-term hernia at the level of L4/5 or L5/S1, in half of cases associated with reactive spondylitis of adjacent vertebral bodies. A significant relationship between Pfirrmann radiological classification of DDD and Thompson morphological assessment system according to the literature data allowed us to use both assessment systems of IVD lesions in patients in our work [10].

We analyzed the expression level of IL-1β, IL-6, IL-17, vascular endothelial growth factor A and CD31 of the removed patients IVD tissue, and compared it with tissue samples of the group without BP. It is important that the expression level of all studied cytokines was positively correlated with the histological degeneration level. This does not leave any doubt about the pathogenetic linkage between immune inflammation and severe degeneration, and also shown the cytokines primacy in the progression of structural disc disorganization [11].

The high expression of IL-1β, IL-6 и IL-17 in the chondroblasts clusters indicates the transformation of the immunomorphogenetic properties of the structural cells of NP and AF, which at one point begin to acquire immunogenic properties. The spatial synergism of IL-1β and IL-17 sheds light on their combined effect in disc degeneration, and allows us to consider their participation in reactive spondylitis and erosion of the vertebral bodies as the main promoters of bone remodeling in DDD by analogy with autoimmune rheumatic diseases (rheumatoid arthritis, psoriatic arthritis etc.) [12, 13].

A high VEGF-A expression level in the NP, which is normally a non-vascular structure, proves the angiogenic abilities of chondrocytes were long before the appearance of a vascular microgrid there. CD31 expression not only in the vascular endothelium, but also in the disc matrix, where vessels were absent, may indicate an increase in the permeability of the disc, since the marker is expressed on cells of the immune system. These data suggest angiogenesis in the discs not so much due to the vessels ingrowth from outside the AF as rather due to their appearance of endotheliocytes in situ, which is confirmed by the data of previous studies in which chondrocytes expressed VEGF-A, VEGF-B and VEGF-C [14]. Thus, local VEGF inhibition at the onset of DDD (2-3 stages by Pfir.) may be considered as a potential target for inhibiting disc degeneration [15]. In our samples, the cracks of AF and NP were not stained with CD31, which denotes their mechanical genesis, not angiogenic, despite the blood cells detected in the lumen. This confirms the assumption of a transformation of the morphogenetic properties of chondroblasts in IVD under the influence of altered disc biomechanics. It is most likely that the start of inflammation in the discs can be caused by hyperphysiological stresses. The relationship between mechanical stress and inflammation in the peak load regions, as previously shown, is realized at the molecular level through the channels of transient receptor potential (TRP) that exist in degenerated IVD. Thus, a locally altered load distribution can potentially affect the morphogenetic properties of fibroblasts through mechanotransduction [3].

EP’s involvement in the process of inflammation according to both histological study and IHC demonstrates the loss of immune privilege by the disc, the spread of immune inflammation beyond its limits to the adjacent areas of the vertebral bodies and the formation of reactive spondylitis and erosive lesions of the bone EP part [16], recorded on MRI images in patients with DDD and BP. So, Ma Kh. L. and co-authors revealed the expression of the same proinflammatory cytokines in vertebral bone marrow with Modic-1 as in the degenerated disc: IL-1, -6 and -8, -10; TNF-α, prostaglandin-E 2, chemotactic protein of monocytes-1, which demonstrates the prevalence of immune inflammation not limited to the border of the disc [17]. A cascade of immune inflammation penetrates disc from the bone marrow of the vertebral bodies through damaged EP. The probability of an inflammatory reaction in the bone in situ is low due to the fact that Modic-changes are detected in patients exclusively in the presence of hernia and/or erosion of the EP, which has been demonstrated in similar studies [18,19]. In turn, the feedback of the EP bone part with the state of the NP cells, their activity and influence on the hydrophilic properties of the extracellular matrix has been demonstrated in studies on the nutritional function of the EP [20].

According to the literature, IL-1β significantly increased the expression of IL-6 and IL-8, VEGF, chemotactic factor (MCP-1) and disk degradation factor (MMP-3) in IVD cells during hypoxia [21]. The level of IL-6 was elevated both in the patients’ serum with DDD and spinal injuries, but the immunohistochemical expression of IL-6 was significantly higher in the degenerative discs tissue [22].

In addition, IL-6 gene increased expression in patients’ blood serum with BP and DDD correlated with inflammatory edema of the bone marrow of adjacent vertebral bodies (Modic-1) compared with individuals who had fatty transformation of vertebral bodies (Modic-2) [23], which point at an inflammatory pattern for all the vertebral-motor segment, and not just for the IVD. A “degenerative profile” with increased expression of VEGF, MMP-2 and MMP-3 was observed in elderly people in all parts of the spine and exceeded that in patients younger than 35 years [24]. VEGF was higher expressed in the elderly in all departments, and the level of IL-1β was higher in the lumbar discs, which does not contradict the results of our study.

Finally, the data of our study allow us to speak about the active immunomorphogenesis of chondrocytes as one of the key factors in the initiation and/or maintenance of disc degeneration. It is known that NP cells may have the properties of stem cells or progenitor cells in the IVD tissue [25] and also the properties of the notochordal cell subpopulation of chondroblasts characteristic for embryonic period of IVD formation [26, 27]. A recent study the heterogeneity of the phenotypes and functions of NP cells (using single-cell RNA sequencing of 5 patients) demonstrated the presence of 3 types of NP cells – chondrocytes (the vast majority of cells with 3 populations – cartilage progenitor cells, fibrochondrocyte progenitor cells and homeostatic chondrocytes), endotheliocytes and macrophages [28]. Based on the marker gene expression specific to the cell line, 7 subclusters were identified among chondrocytes, differing in heterogeneity and biological functions. Thus, the C-1 and -3 subclusters of chondrocytes had an inflammatory phenotype. It is these cell lines that can be considerably activated during disc degeneration, which explains the transformation phenomenon of the NP cells immunomorphogenesis at the level of the entire population of chondrocytes, i.e. their pathological differentiation in DDD. Moreover, inflammatory subclusters of chondrocytes and macrophages, which presents in much smaller numbers in the NP, another subcluster is able to synthesize the angiogenesis factor and enhance the adhesion of endotheliocytes. Thus, individual immune phenotypes of degenerated disc chondrocytes act as conductors of the inflammatory cascade and vascularization in IVD.

In this study, low levels of expression of cytokines IL-1β, -6, -17, and VEGF-A were observed in the samples of the control group, which allows for the probability of the onset of disc «predegeneration», which did not manifest histologically and at the macro level (no signs of degeneration were detected visually and microscopically).

This finding may indicate an early preclinical and pre-morphological stage of IVD degeneration, which can manifest itself by a displaced cytokine profile and altered mechanoelastic disc functions with pronounced axial loads, which requires further study.

## CONCLUSIONS

Our data from clinical and instrumental studies of patients with BP and DDD in combination with histological and immunohistochemical analysis of degenerated IVD tissues provided a three-dimensional picture of the “inflammatory-degenerative pattern” in the lumbar spine degeneration and the formation of IVD hernia. Immune inflammation in discs with IL-1β, -6, -17 was present in all discs of the DDD group (p < 0.0001) and had a high level of correlation with the histological stage of IVD degeneration (r > 0.5; p < 0.0001). This proves that local immune inflammation is an obligatory component of the degenerative cascade in the IVD, initiates angiogenesis in the NP, AF of discs with the expression of angiogenesis factors VEGF-A on the surface of chondrocytes and CD31 in the extracellular matrix, and extends to the EP. It is a key mechanism for the development of reactive osteitis of adjacent vertebrae with formation of inflammatory erosive lesions of the vertebral-motor segment in BP. The detection of cytokine expression on the surface of chondrocytes, including clusters of chondroblasts, demonstrates altered morphogenetic properties of chondrocytes with transformation into an inflammatory cell phenotype and the secondary role of macrophages in IVD tissue. The results obtained will help to identify molecular and cellular targets and develop basic strategies for biological, cellular and targeted therapy of BP in young people at the early stages of DDD.

## LIMITATIONS OF THE STUDY

The study included only those patients who were shown surgery due to the ineffectiveness of conservative therapy, which causes a shift in clinical and morphological indicators towards more severe data. We could not assess the disc degeneration by MRI in a group of healthy donors, conclusions about its absence or minimal manifestations are based only on the data of the pathomorphological findings.

## Data Availability

All data produced in the present study are available upon reasonable request to the authors.

## Acknowledgements

head the educational department of the Pirogov Medical University Pirogov, doctor of medical sciences prof. Aksenova A.V., neurosurgeons of the Pirogov City Clinical Hospital No. 1, M. A. Nekrasov, V. V. Babenkov, A. N. Isayev, V. A. Smirnov, D. S. Glukhov, G. V. Gabechiya, D. B. Choriyev, A. Kh. Kozhev for help in the research.

## Contribution of the authors

N. G. Pravdyuk – literature analysis, problem statement, research design development, data analysis and interpretation; A.V. Novikova – literature analysis, collection of clinical data and a set of biomaterials, performance of the preanalytical stage of work, analysis and interpretation of data, preparation of drawings and graphs; N. A. Shostak – research management, research design development, data interpretation, manuscript editing; A.A. Buianova – literature analysis, biomaterial set, performing analytical part of laboratory work, determination and statistical analysis of the expression level of immunohistochemical markers in biomaterial, preparation of drawings and graphs; O. I. Patsap – admission to work with biological material in the department of pathological anatomy, performing the analytical part of laboratory work; A. P. Raksha – admission to work with biological material; V. M. Feniksov, D. A. Nikolayev, I.V. Senko – admission to a set of clinical and biological material during discectomy, neurological examination of patients; V. T. Timofeyev – planning the study, performing the preanalytical stage of the work, evaluating the data obtained.

## REFERENCES

1. Cheng, Chuan, et al. “Lactylation driven by lactate metabolism in the disc accelerates intervertebral disc degeneration: A hypothesis.” Medical Hypotheses 159 (2022): 110758.

2. Boos, Norbert et al. “Classification of age-related changes in lumbar intervertebral discs: 2002 Volvo Award in basic science.” Spine vol. 27,23 (2002): 2631–44. doi:10.1097/00007632-200212010-00002

3. Sadowska, Aleksandra, et al. “Inflammaging in the Intervertebral Disc.” Clinical and Translational Neuroscience, vol. 2, no. 1, Jan. 2018, p. 2514183X1876114. Crossref, https://doi.org/10.1177/2514183X18761146.

4. Risbud, Makarand V, and Irving M Shapiro. “Role of cytokines in intervertebral disc degeneration: pain and disc content.” Nature reviews. Rheumatology vol. 10,1 (2014): 44–56. doi:10.1038/nrrheum.2013.160

5. Schnake, Klaus John et al. “Mechanical concepts for disc regeneration.” European spine journal : official publication of the European Spine Society, the European Spinal Deformity Society, and the European Section of the Cervical Spine Research Society vol. 15 Suppl 3,Suppl 3 (2006): S354–60. doi:10.1007/s00586-006-0176-y

6. Kos, Natasa et al. “A Brief Review of the Degenerative Intervertebral Disc Disease.” Medical archives (Sarajevo, Bosnia and Herzegovina) vol. 73,6 (2019): 421–424. doi:10.5455/medarh.2019.73.421-424

7. De Geer, Christopher M. “Cytokine Involvement in Biological Inflammation Related to Degenerative Disorders of the Intervertebral Disk: A Narrative Review.” Journal of chiropractic medicine vol. 17,1 (2018): 54–62. doi:10.1016/j.jcm.2017.09.003

8. Molinos, Maria et al. “Inflammation in intervertebral disc degeneration and regeneration.” Journal of the Royal Society, Interface vol. 12,104 (2015): 20141191. doi:10.1098/rsif.2014.1191

9. Sive, J I et al. “Expression of chondrocyte markers by cells of normal and degenerate intervertebral discs.” Molecular pathology : MP vol. 55,2 (2002): 91–7. doi:10.1136/mp.55.2.91

10. Pękala, P et al. “Correlation of morphological and radiological characteristics of degenerative disc disease in lumbar spine: a cadaveric study.” Folia morphologica vol. 81,2 (2022): 503–509. doi:10.5603/FM.a2021.0040

11. Paesold, Günther et al. “Biological treatment strategies for disc degeneration: potentials and shortcomings.” European spine journal : official publication of the European Spine Society, the European Spinal Deformity Society, and the European Section of the Cervical Spine Research Society vol. 16,4 (2007): 447–68. doi:10.1007/s00586-006-0220-y

12. Chabaud, M et al. “Enhancing effect of IL-17 on IL-1-induced IL-6 and leukemia inhibitory factor production by rheumatoid arthritis synoviocytes and its regulation by Th2 cytokines.” Journal of immunology (Baltimore, Md. : 1950) vol. 161,1 (1998): 409–14.

13. Blauvelt, Andrew, and Andrea Chiricozzi. “The Immunologic Role of IL-17 in Psoriasis and Psoriatic Arthritis Pathogenesis.” Clinical reviews in allergy & immunology vol. 55,3 (2018): 379–390. doi:10.1007/s12016-018-8702-3

14. Dai, J, and A B M Rabie. “VEGF: an essential mediator of both angiogenesis and endochondral ossification.” Journal of dental research vol. 86,10 (2007): 937–50. doi:10.1177/154405910708601006

15. Scholz, B et al. “Suppression of adverse angiogenesis in an albumin-based hydrogel for articular cartilage and intervertebral disc regeneration.” European cells & materials vol. 20 24–36; discussion 36-7. 13 Jul. 2010, doi:10.22203/ecm.v020a03

16. Xiong, Chengjie et al. “Migration inhibitory factor enhances inflammation via CD74 in cartilage end plates with Modic type 1 changes on MRI.” Clinical orthopaedics and related research vol. 472,6 (2014): 1943–54. doi:10.1007/s11999-014-3508-y

17. Ma, Xin-Long et al. “Possible role of autoimmune reaction in Modic Type I changes.” Medical hypotheses vol. 76,5 (2011): 692–4. doi:10.1016/j.mehy.2011.01.035

18. Lotz, J C et al. “The role of the vertebral end plate in low back pain.” Global spine journal vol. 3,3 (2013): 153–64. doi:10.1055/s-0033-1347298

19. Novikova A V et al. “DEGENERATIVE DISC DISEASE IN YOUNG ADULTS: CYTOKINE PROFILE AND ANGIOGENIC FACTORS.” Bulletin of Russian State Medical University, no. 6, (2021): 75–83. doi:10.24075/vrgmu.2021.061

20. Fields, Aaron J et al. “Contribution of the endplates to disc degeneration.” Current molecular biology reports vol. 4,4 (2018): 151–160. doi:10.1007/s40610-018-0105-y

21. Hsu, Yu-Hsiang et al. “Effects of IL-1β, IL-20, and BMP-2 on Intervertebral Disc Inflammation under Hypoxia.” Journal of clinical medicine vol. 9,1 140. 4 Jan. 2020, doi:10.3390/jcm9010140

22. Rodrigues, Luciano Miller Reis et al. “Inflammatory biomarkers in sera of patients with intervertebral disc degeneration.” Einstein (Sao Paulo, Brazil) vol. 17,4 eAO4637. 29 Aug. 2019, doi:10.31744/einstein_journal/2019AO4637

23. Schroeder, Gregory D et al. “Are Modic changes associated with intervertebral disc cytokine profiles?” The spine journal: official journal of the North American Spine Society vol. 17,1 (2017): 129–134. doi:10.1016/j.spinee.2016.08.006

24. Baptista, Josemberg S et al. “Expression of degenerative markers in intervertebral discs of young and elderly asymptomatic individuals.” PloS one vol. 15,1 e0228155. 27 Jan. 2020, doi:10.1371/journal.pone.0228155

25. Risbud, Makarand V et al. “Evidence for skeletal progenitor cells in the degenerate human intervertebral disc.” Spine vol. 32,23 (2007): 2537–44. doi:10.1097/BRS.0b013e318158dea6

26. Gilson, Audrey et al. “Differential expression level of cytokeratin 8 in cells of the bovine nucleus pulposus complicates the search for specific intervertebral disc cell markers.” Arthritis research & therapy vol. 12,1 (2010): R24. doi:10.1186/ar2931

27. Lawson, Lisa, and Brian D Harfe. “Notochord to Nucleus Pulposus Transition.” Current osteoporosis reports vol. 13,5 (2015): 336–41. doi:10.1007/s11914-015-0284-x

28. Han, Shuo et al. “Single-Cell RNA Sequencing of the Nucleus Pulposus Reveals Chondrocyte Differentiation and Regulation in Intervertebral Disc Degeneration.” Frontiers in cell and developmental biology vol. 10 824771. 21 Feb. 2022, doi:10.3389/fcell.2022.824771

